# Epidemiological and Clinical Characteristics of Coronavirus Disease 2019 in Shenzhen, the Largest Migrant City of China

**DOI:** 10.1101/2020.03.22.20035246

**Authors:** Ying Wen, Lan Wei, Yuan Li, Xiujuan Tang, Shuo Feng, Kathy Leung, Xiaoliang Wu, Xiong-Fei Pan, Cong Chen, Junjie Xia, Xuan Zou, Tiejian Feng, Shujiang Mei

## Abstract

We conducted a retrospective study among 417 confirmed COVID-19 cases from Jan 1 to Feb 28, 2020 in Shenzhen, the largest migrant city of China, to identify the epidemiological and clinical features in settings of high population mobility. We estimated the median incubation time to be 5.0 days. 342 (82.0%) cases were imported, 161 (38.6%) cases were identified by surveillance, and 247 (59.2%) cases were reported from cluster events. The main symptoms on admission were fever and dry cough. Most patients (91.4%) had mild or moderate illnesses. Age of 50 years or older, breathing problems, diarrhea, and longer time between the first medical visit and admission were associated with higher level of clinical severity. Surveillance-identified cases were much less likely to progress to severe illness. Although the COVID-19 epidemic has been contained in Shenzhen, close monitoring and risk assessments are imperative for prevention and control of COVID-19 in future.

**Article Summary Line:** We characterized epidemiological and clinical features of a large population-based sample of COVID-19 cases in the largest migrant city of China, and our findings could provide knowledge of SARS-CoV-2 transmission in the context of comprehensive containment and mitigation efforts in similar settings.

## Introduction

The Coronavirus Disease 2019 (COVID-19) caused by Severe Acute Respiratory Syndrome Coronavirus 2 (SARS-CoV-2) is emerging as a major infectious disease epidemic globally. Initially detected in a cluster of patients with unexplained pneumonia in Wuhan, Hubei Province of China in early December 2019, SARS-CoV-2 rapidly spread not only within China but also around the globe within just three months. As of Mar 10, 2020, 113,702 confirmed cases and 4,012 deaths have been reported in 109 countries (*1*). Although the natural reservoir of SARS-CoV-2 is still unknown, early confirmed cases are strongly associated with exposures to wild animals in the Huanan Seafood Wholesale Market (*2*), and sustained human-to-human transmission is observed particularly among close contacts (*3, 4*). Due to the surging number and rapid spread of COVID-19, World Health Organization (WHO) has increased the risk assessment of COVID-19 to ‘very high’ at the global level on Feb 28, 2020.

As one of the most developed and commercialized cities in China, Shenzhen is the largest migrant city where over 80% of its population (20 million) are migrants. The risk of case importation in Shenzhen was therefore high, especially because the COVID-19 epidemic occurred around the Chinese Lunar New Year holiday season during which intra-city mobility of the migrant population was extremely high. Since the first case of COVID-19 in Shenzhen was confirmed on Jan 20, 2020, local authorities activated the highest level of emergency response to the disease. Although there have been several studies on the transmission, epidemiology and clinical symptoms of COVID-19 in Wuhan (*3, 5, 6*), data from other epidemic areas are still lacking (*7*). In particular, information and knowledge of COVID-19 from a migrant city with high population mobility like Shenzhen can inform effective prevention and control strategies in other similar settings. As such, we investigated the epidemiological and clinical characteristics of all 417 cases that were confirmed in Shenzhen as of Feb 28, 2020.

## Methods

### Data Collection

We conducted a retrospective study of the epidemiological and clinical characteristics of COVID-19 cases in Shenzhen from Jan 1 to Feb 28, 2020. COVID-19 case was defined in accordance with the WHO interim guidelines and the National Guidelines in Diagnosis and Treatment Scheme for COVID-19 (Sixth edition) (*8, 9*). Sputum, blood, broncho-alveolar lavage fluid, nasopharyngeal swab, or oropharyngeal swab were collected from each patient and tested using real-time polymerase chain reaction (RT-PCR) for SARS-CoV-2. A cycle threshold value less than 37 was defined as PCR test positive. Virus detection was first done by hospitals or districted centers for disease control and prevention (CDC), and positive results were further confirmed by Guangdong Provincial CDC (before Jan 30, 2020) or Shenzhen CDC (after Jan 30, 2020). For each confirmed case, detailed epidemiological investigations were conducted by local epidemiologists and public health workers. All confirmed cases were immediately reported to the National Infectious Disease Information System for COVID-19, which was amended as a Class B notifiable infectious disease on Jan 20, 2020.

We compiled the epidemiological and clinical data from both local epidemiological investigation reports and the National Infectious Disease Information System. Patients’ information including sociodemographic characteristics, exposure history, close contacts, time-lines of illness onset, medical visit, hospitalization, and PCR confirmation, symptoms, and clinical outcomes, was extracted to construct a dataset with no personal identity. All confirmed COVID-19 cases in Shenzhen were included in the current analyses with no pre-specified exclusion criteria. Data collection in the epidemiological investigation was part of the continuing public health investigation of an emerging outbreak and therefore the individual informed consent was waived. The study was approved by the ethics committees of Shenzhen Center for Disease Control and Prevention.

### Study definition

Patients were defined as having Wuhan exposure if they were Wuhan residents or visited Wuhan within the past 14 days before symptom onset, and were defined as having Shenzhen exposure if they had not left Shenzhen within the past 14 days before symptom onset, while all others exposed elsewhere were defined as having exposure elsewhere in mainland China other than Wuhan or Shenzhen. Based on the National Guidelines in Diagnosis and Treatment Scheme for COVID-19 (*9*), the seriousness of clinical presentations of COVID-19 cases was categorized as mild, moderate, severe, and critical. Mild cases were those with virological confirmation but without an evidence of having pneumonia. Moderate cases were mild cases at the same time with a diagnosis of pneumonia. Severe was defined when one of the following criteria was met: dyspnea (respiratory frequency≥30/minute); blood oxygen saturation ≤93%; PaO2/FiO2 ratio <300; and/or lung infiltrates >50% within 24–48 hours.

Critical cases were defined when patients had respiratory failure; and/or septic shock; and/or multiple organ dysfunction/failure. In addition, we categorized cases into self-identified and surveillance-identified ones. The former referred to COVID-19 cases who were identified when they sought medical care at the hospitals, while the latter referred to cases who were identified through active surveillance efforts including screening of close contacts of the confirmed patients and recent travelers from Hubei; fever monitoring at airport, train station, docks, and highway checkpoints; and registration and report of fever by community workers. We defined a cluster as occurrence of two or more confirmed cases in a socially-close setting (such as a family, a school, or a company) within the past 14 days, which may be caused by human-to-human transmission through close contacts within such setting or infection via a common external exposure. Other cases not from a cluster were defined as scattered cases.

### Statistical analysis

Demographic and clinical characteristics as categorical variables were presented as numbers and percentages, while continuous variables were presented as means and standard deviations or 95% confidence intervals (CI), or medians and interquartile ranges (IQR) if appropriate. Inter-group differences in the characteristics were tested by using Pearson’s χ^2^ test or Fisher’s exact test for categorical variables, and by using Student’s t-test or analysis of variance for continuous variables showing a normal distribution, and Kruskal–Wallis and Wilcoxon tests for continuous variables with non-parametric distribution. The incubation period was estimated by using a previously described parametric accelerated failure time model (*10*). Patients with detailed information on the time of exposure, the date of illness onset, or the first time of presentation were included for this analysis. We fitted lognormal, gamma, and Weilbull distributions using Markov Chain Monto Carlo in a Bayesian framework (*11*). We estimated the serial interval by using the time difference of illness onset between the infector and infectee. Initial reproductive number was estimated by using the best fit model based on date of illness onset of the early (Jan 10-23) local exposed cases without relation to the imported cases and the estimated serial interval of COVID-19. Logistic regression models were applied to identify factors associated with the clinical severity of COVID-19. All statistical tests and analyses of the incubation period, serial interval, and initial reproductive number were performed in R software (R foundation for Statistical Computing).

## Results

A total of 417 cases had been confirmed as of Feb 28, 2020 in Shenzhen. In terms of potential source of exposure, 224 (53.7%) were exposed in Wuhan, 75 (18.0%) were exposed in Shenzhen, and 118 (28.3%) were exposed elsewhere (Table 1). In addition, 161 (38.6%) cases were identified by surveillance, and 247 (59.2%) cases occurred in 92 clusters (common exposure or secondary transmission). None of the patients had been to the Huanan Seafood Wholesale Market within the past 14 days before illness onset, which was initially thought to be the index location of zoonotic infections of SARS-CoV-2 that started the COVID-19 epidemic (*12*).

**Table 1.**
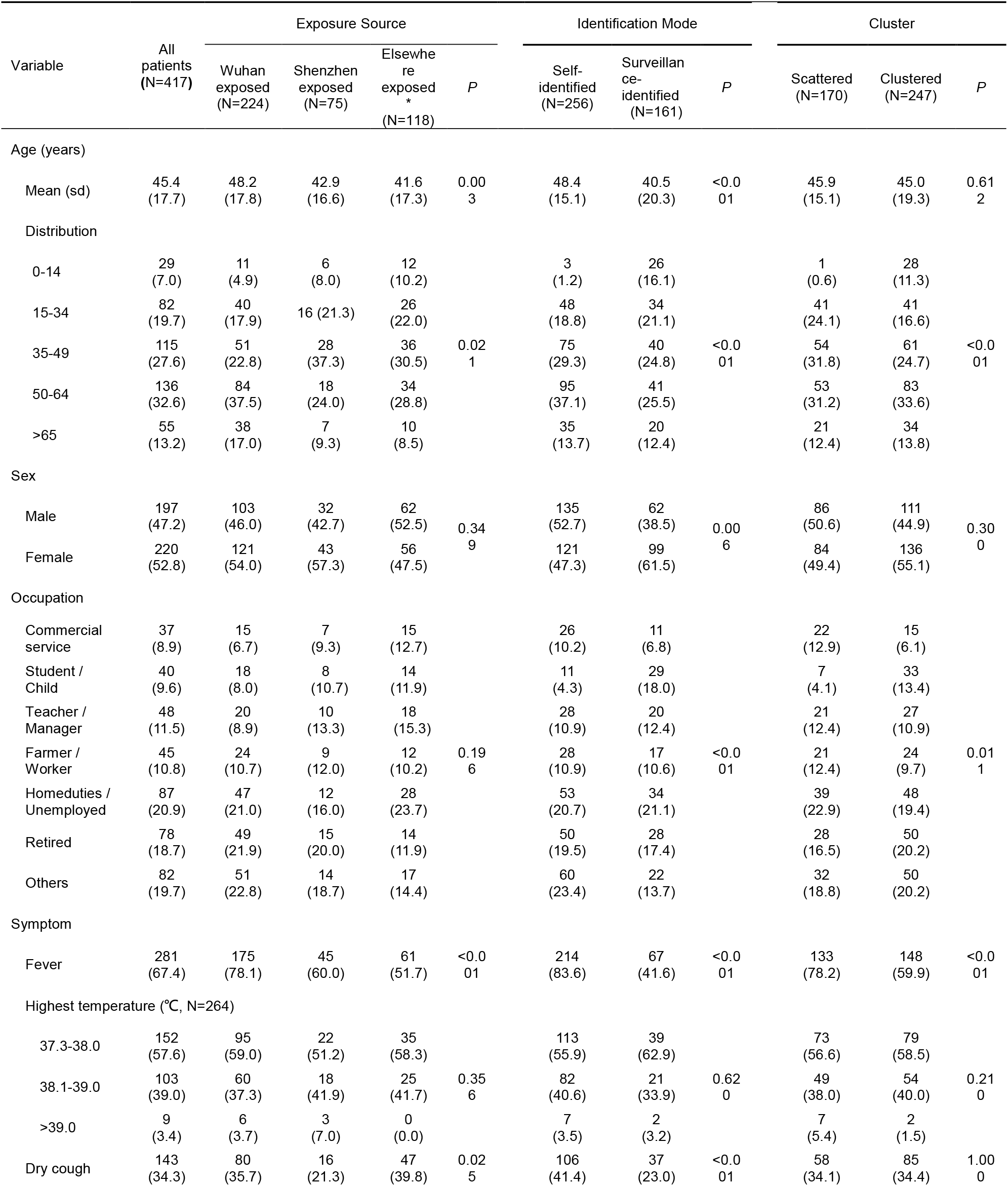
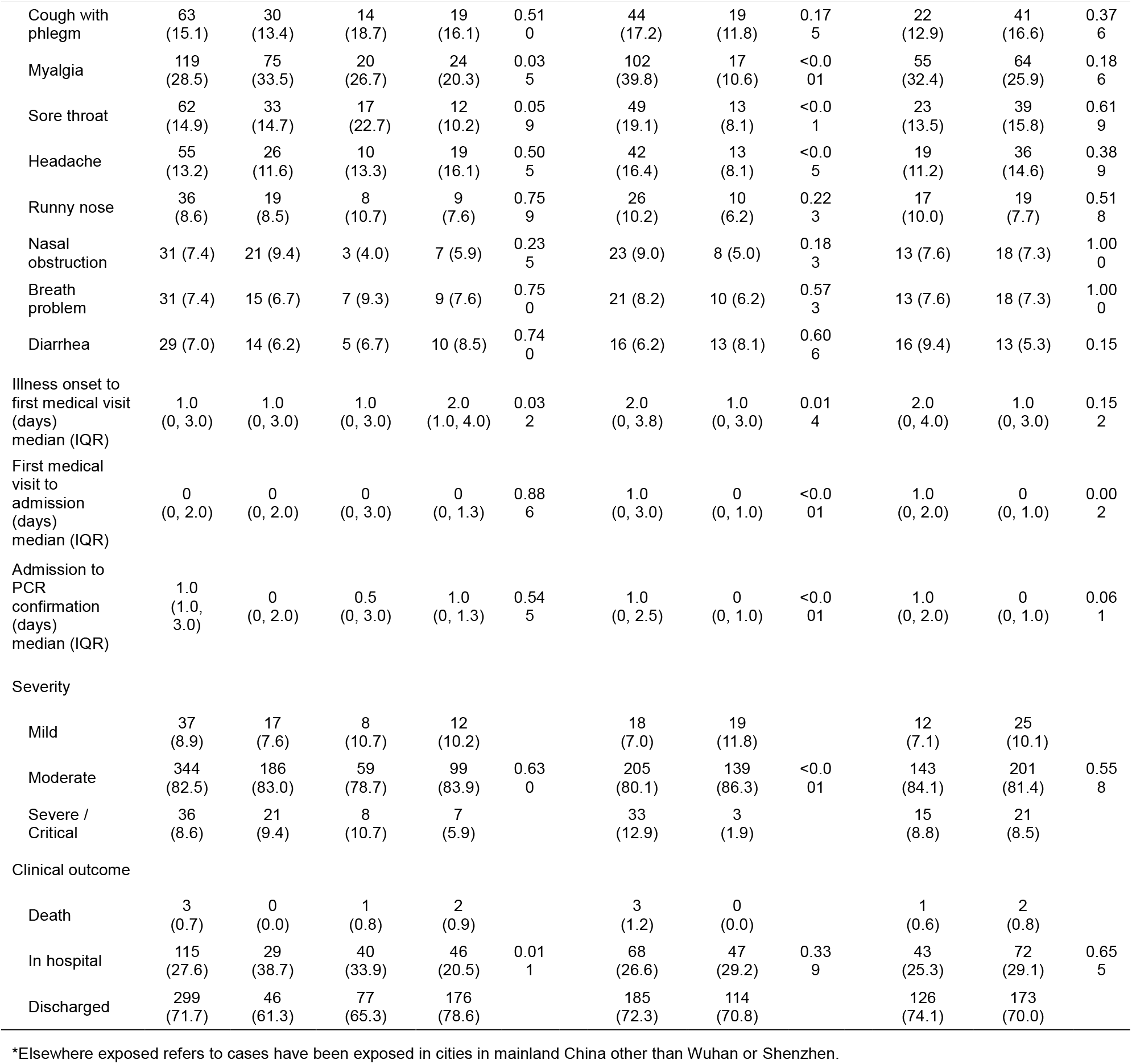
Demographics and clinical symptoms of Coronavirus Disease 2019 cases

Figure 1 showed the epidemic curve of COVID-19 cases by the date of illness onset and PCR confirmation against the timeline of a series of public health responses, policies and intervention measures. The number of COVID-19 cases in Shenzhen increased slowly in the early stage (Jan 1-15), then experienced a short rapid growth phase until Jan 23, 2020 and reached a plateau, and finally showed a sustained downward trend. Regarding the source of infection, the cases were more likely to have Wuhan exposure if they had illness onset before the Chinese Lunar New Year holiday starting from Jan 24, while more cases had exposure in Shenzhen or elsewhere afterwards (Figure 2A). In addition, there were more cases who sought medical care after illness onset in the early phase, while cases were more likely to be identified by surveillance and to be reported from cluster events after Jan 20 (Figure 2B, 2C).

**Figure 1.**
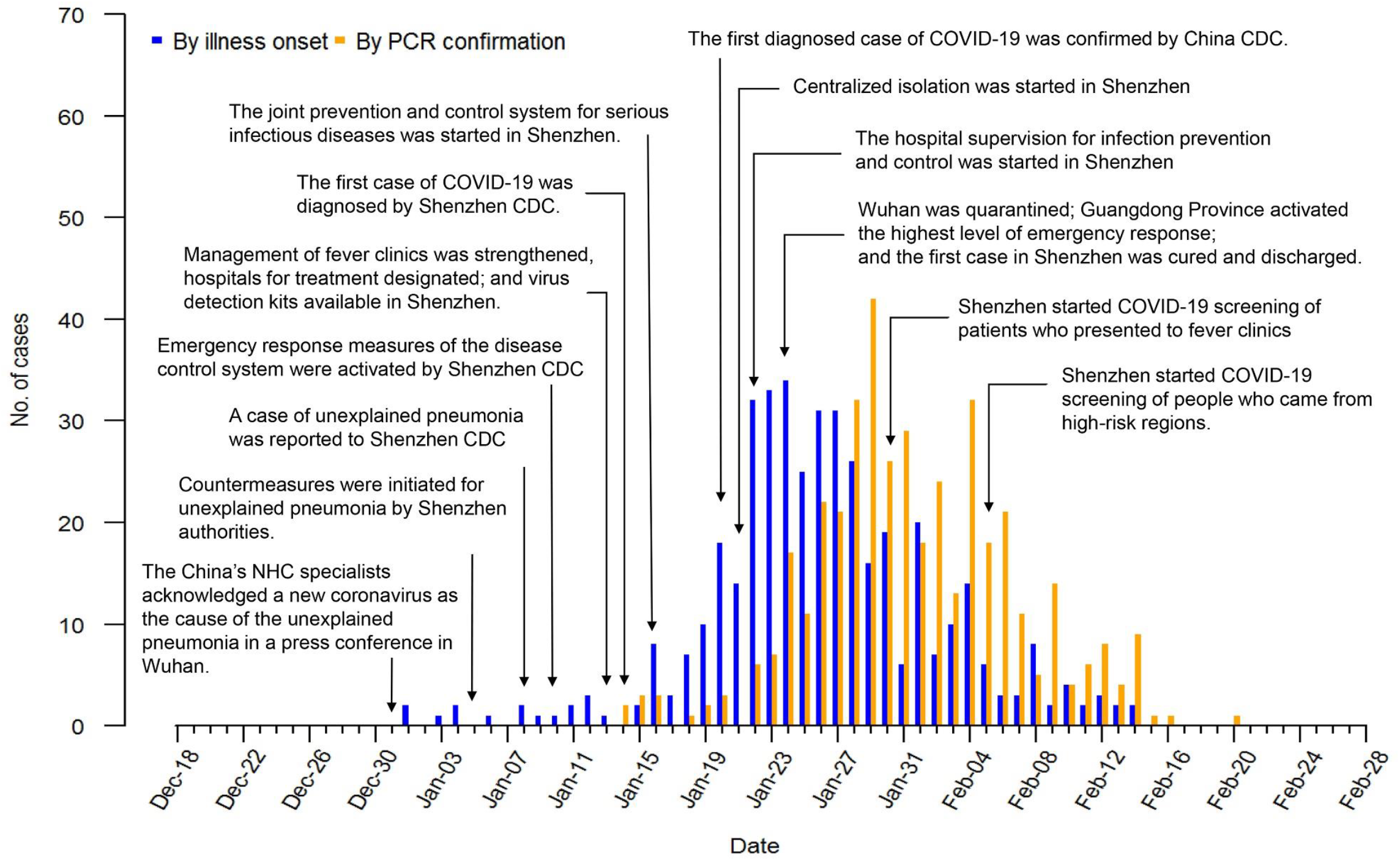
The epidemic curve of COVID-19 cases by the date of illness onset and PCR confirmation in Shenzhen.

**Figure 2.**
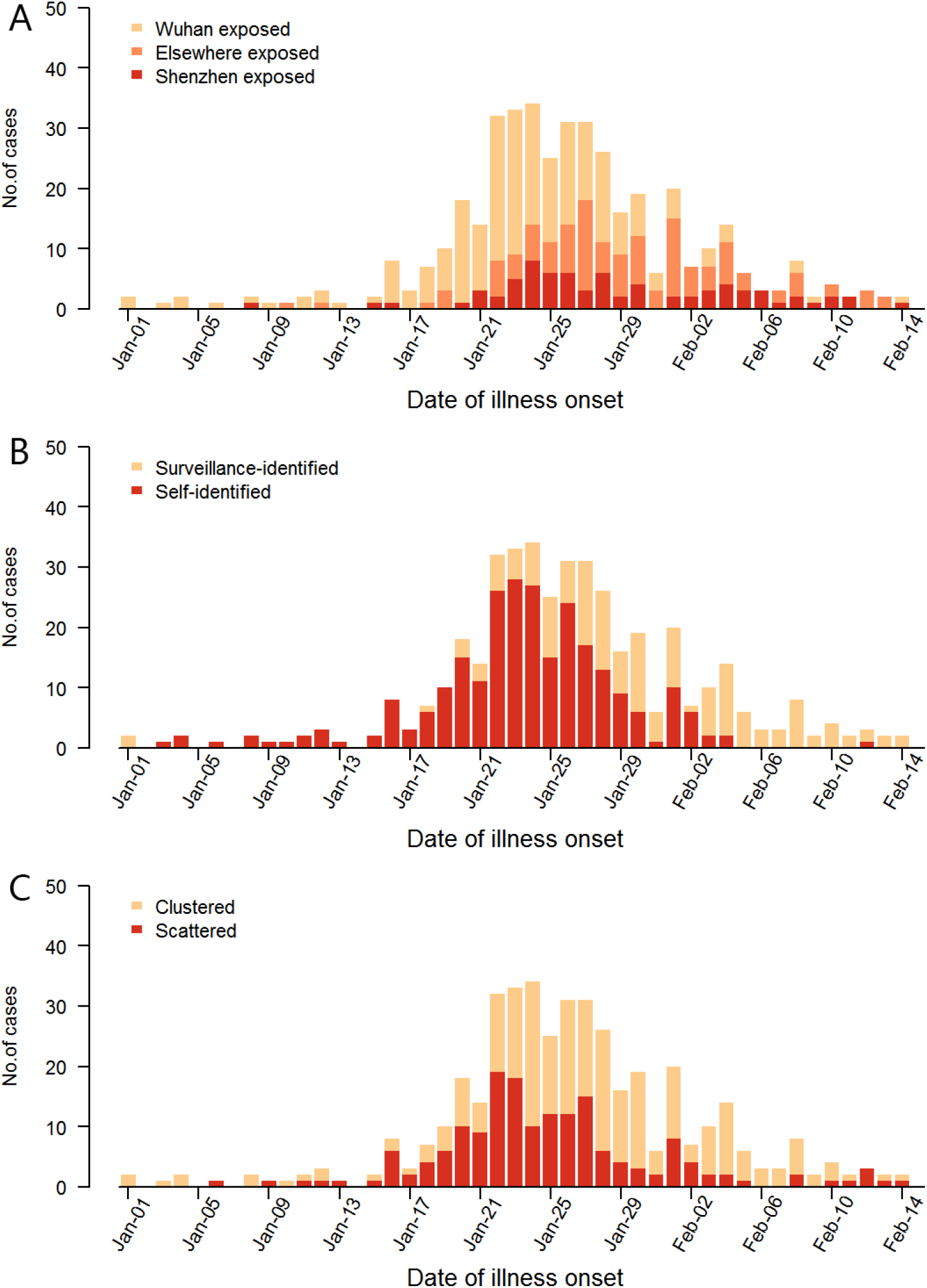
The confirmed COVID-19 cases by the date of illness onset of different groups. A) exposure source. B) identification mode. C) cluster.

Using detailed information on the time of exposure and illness onset from 92 patients, we estimated the median incubation time of SARS-CoV-2 infection to be 5.0 (IQR: 3.1-8.2) days assuming the incubation time followed a lognormal distribution. Estimates from models with other distributions (gamma and Weilbull) were 5.4 to 5.5 days (See supplemental materials). Using information on the date of illness onset from 28 pairs of infector and infectee, we estimated the mean serial interval to be 5.5 (95% CI: 4.1-7.0) days. We estimated the initial reproductive number to be 2.5 (95% CI: 1.4-4.3) using information from the cases that exposed in Shenzhen.

Demographic characteristics, clinical symptoms and outcomes were presented in Table 1. The mean age was 45.4 years old, and 220 (52.8%) cases were female. Patients with Wuhan exposure were older than those with exposure in Shenzhen or elsewhere (48.2 vs 42.9, 41.6, *P*<0.05). Patients identified by surveillance were younger than those self-identified cases (40.5 vs 48.4, *P*<0.001). In total, there were 29 (7.0%) pediatric patients aged below 15 years, of whom 26 were identified by surveillance and 28 were cases from clusters.

The most common symptoms of COVID-19 were fever (67.4%), dry cough (34.3%), and myalgia (28.5%). Patients who were exposed in Wuhan (78.1%), identified by surveillance (83.6%), or scattered (78.2%) were more likely to have fever (*P*<0.001). Patients with Shenzhen exposure were less likely to have dry cough (21.3%) than patients with Wuhan exposure (35.7%) and elsewhere exposure (39.8%); they were also less likely to have myalgia (26.7%) than those with Wuhan exposure (33.5%). Patients who were identified by surveillance were less likely to have symptoms than the self-identified, such as fever (41.6% vs 83.6%), dry cough (23.0% vs 41.4%), myalgia (10.6% vs 39.8%), sore throat (8.1% vs 19.1%), and headache (8.1% vs 16.4%).

The median time interval was 1.0 (IQR: 0-3.0) day from illness onset to the first medical visit, 0 (IQR: 0-2.0) day from the first medical visit to hospital admission, and 1.0 (IQR: 1.0-3.0) day from hospital admission to PCR confirmation (Table 1). Patients with elsewhere exposure showed a longer time interval from illness onset to the first medical visit (2.0, IQR: 1.0-4.0) than those with Shenzhen exposure (1.0, IQR: 0-3.0) or Wuhan exposure (1.0, IQR: 0-3.0) (*P*< 0.05; Table 1, Figure 3A). Generally, surveillance-identified patients had shorter time intervals between illness onset, the first medical visit, hospital admission, and PCR confirmation than those self-identified patients, with the estimates to be 1.0 vs 2.0 days, 0 vs 1.0 day, 0 vs 1.0 day, respectively (*P*<0.05 for all; Table 1, Figure 3B). The scattered cases had a longer time interval from the first medical visit to hospital admission than the clustered cases (1.0 vs 0 day, *P*<0.05; Table 1, Figure 3C).

**Figure 3.**
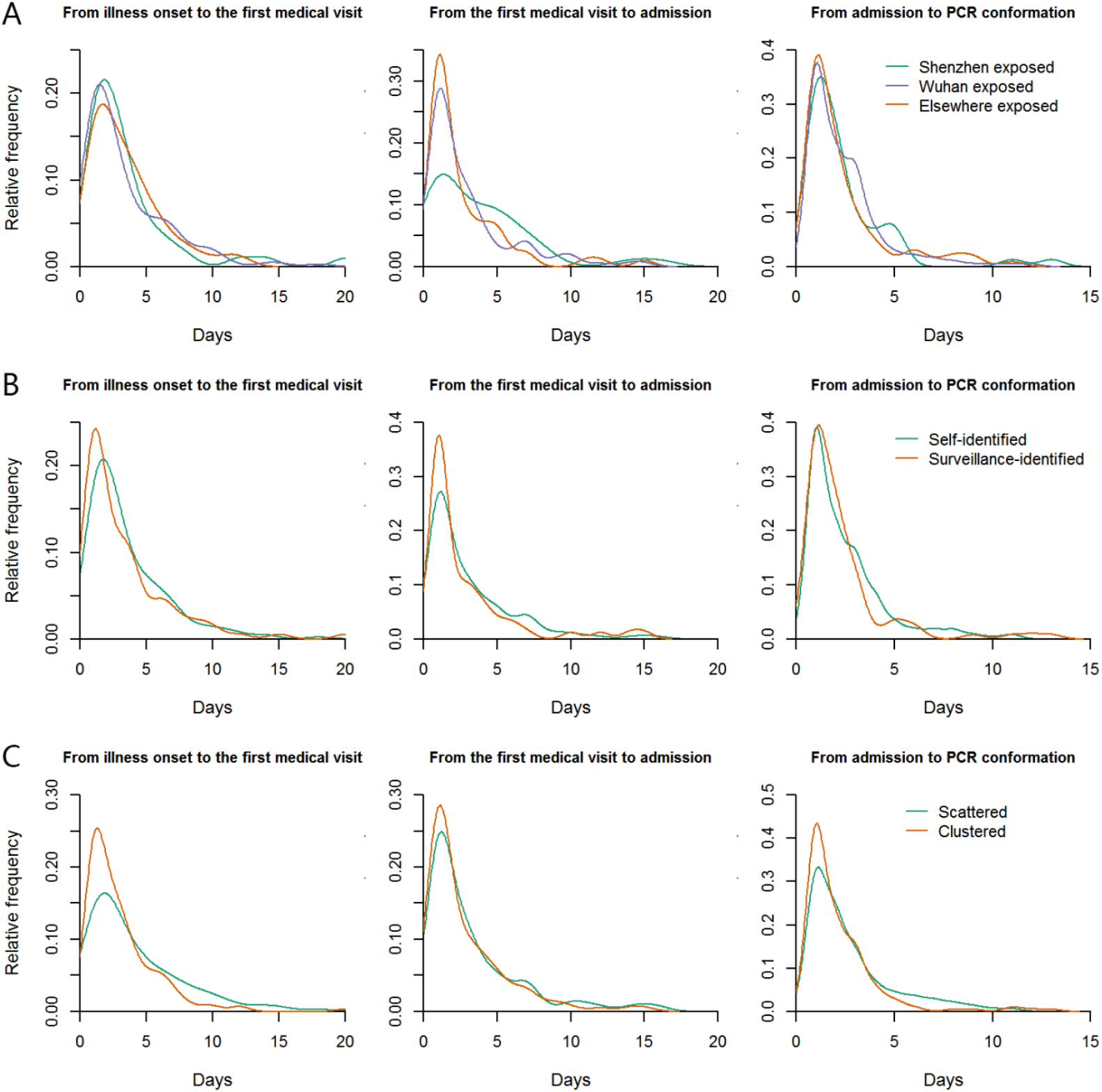
Key time-to-event distribution of different groups. A) exposure source. B) identification mode. C) cluster.

Most patients had mild (37; 8.9%) or moderate (344; 82.5%) conditions on admission, and only 36 (8.6%) were severe or critical cases (30 severe pneumonia and 6 critical pneumonia). 83.3% (30/36) of the severe or critical cases occurred in patients aged 50 or above. The clinical severity of patients with different sources of exposure was not significantly different, whereas the self-identified patients had a significantly higher percentage of severe or critical cases than those identified by surveillance (12.9% vs 1.9%, *P*<0.001). As of Feb 28, 299 (71.7%) patients were discharged, 115 (27.6%) patients were under treatment, and 3 (0.7%) patients were dead. The hospital fatality rate was 3/417 (0.7%) in this study.

We tentatively explored factors that correlated with the clinical severity of COVID-19 on admission (Table 2). Age of 50 years or older (adjusted odds ratio, AOR: 6.15; 95% CI: 2.22-17.03), breathing problems (including shortness of breath, dyspnea, or chest tightness) (AOR: 9.60; 95% CI: 3.04-30.34), diarrhea (AOR: 3.65; 95% CI: 1.04-12.74), and longer time between the first medical visit and admission (AOR: 1.20; 95% CI: 1.06-1.36) were associated with higher level of clinical severity. Cases identified by surveillance were much less likely to progress to severe illness (AOR: 0.17; 95% CI: 0.04-0.66).

**Table 2.** Factors associated with severe pneumonia caused by Severe Acute Respiratory Syndrome Coronavirus 2

| Variable | Mild / Moderate<br>(N=381) | Severe / Critical<br>(N=36) | P | OR (95%CI) | AOR (95%CI)* |
| --- | --- | --- | --- | --- | --- |
| Age (years) |  |  |  |  |  |
| <50 | 220 (57.7) | 6 (16.7) | <0.001 | 1.00 | 1.00 |
| ≥50 | 161 (42.3) | 30 (83.3) |  | 6.83 (2.78, 16.80) | 6.15 (2.22, 17.03) |
| Sex |  |  |  |  |  |
| Male | 174 (45.7) | 23 (63.9) | 0.053 | 1.00 | - |
| Female | 207 (54.3) | 13 (36.1) |  | 0.48 (0.23, 0.97) | - |
| Fever |  |  |  |  |  |
| Yes | 248 (65.1) | 33 (91.7) | 0.001 | 5.90 (1.77, 19.62) | 3.04 (0.77, 12.02) |
| No | 133 (34.9) | 3 (8.3) |  | 1.00 | 1.00 |
| Dry cough |  |  |  |  |  |
| Yes | 125 (32.8) | 18 (50.0) | 0.044 | 2.05 (1.03, 4.08) | - |
| No | 256 (67.2) | 18 (50.0) |  | 1.00 | - |
| Cough with phlegm |  |  |  |  |  |
| Yes | 59 (15.5) | 4 (11.1) | 0.629 | 0.68 (0.23, 2.00) | 0.38 (0.11, 1.36) |
| No | 322 (84.5) | 32 (88.9) |  | 1.00 | 1.00 |
| Myalgia |  |  |  |  |  |
| Yes | 102 (26.8) | 17 (47.2) | 0.012 | 2.45 (1.23, 4.89) | - |
| No | 279 (73.2) | 19 (52.8) |  | 1.00 | - |
| Sore throat |  |  |  |  |  |
| Yes | 59 (15.5) | 3 (8.3) | 0.331 | 0.50 (0.15, 1.67) | 0.38 (0.10, 1.47) |
| No | 322 (84.5) | 33 (91.7) |  | 1.00 | 1.00 |
| Headache |  |  |  |  |  |
| Yes | 52 (13.6) | 3 (8.3) | 0.604 | 0.58 (0.17, 1.94) | 0.33 (0.08, 1.47) |
| No | 329 (86.4) | 33 (91.7) |  | 1.00 | 1.00 |
| Running nose |  |  |  |  |  |
| Yes | 34 (8.9) | 2 (5.6) | 0.756 | 0.60 (0.14, 2.61) | - |
| No | 347 (91.1) | 34 (94.4) |  | 1.00 | - |
| Nasal obstruction |  |  |  |  |  |
| Yes | 29 (7.6) | 2 (5.6) | 0.998 | 0.71 (0.16, 3.12) | - |
| No | 352 (92.4) | 34 (94.4) |  | 1.00 | - |
| Breath problem |  |  |  |  |  |
| Yes | 21 (5.5) | 10 (27.8) | <0.001 | 6.59 (2.81, 15.47) | 9.60 (3.04, 30.34) |
| No | 360 (94.5) | 26 (72.2) |  | 1.00 | 1.00 |
| Diarrhea |  |  |  |  |  |
| Yes | 23 (6.0) | 6 (16.7) | 0.029 | 3.11 (1.18, 8.23) | 3.65 (1.04, 12.74) |
| No | 358 (94.0) | 30 (83.3) |  | 1.00 | 1.00 |
| Illness onset to first medical visit (days) Median (IQR) | 1.0 (0, 3.0) | 2.0 (0.8, 4.0) | 0.056 | 1.06 (0.96, 1.16) | 1.13 (0.99, 1.28) |
| First medical visit to admission (days) Median (IQR) | 0 (0, 1.0) | 3.0 (0, 4.3) | <0.001 | 1.18 (1.08, 1.29) | 1.20 (1.06, 1.36) |
| Admission to PCR confirmation (days) Median (IQR) | 1.0 (0, 2.0) | 1.5 (1.0, 3.3) | 0.050 | 1.11 (0.98, 1.27) | - |
| Exposure Source |  |  |  |  |  |
| Shenzhen exposed | 67 (17.6) | 8 (22.2) | 0.419 | 1.00 | - |
| Wuhan exposed | 203 (53.3) | 21 (58.3) |  | 0.87 (0.37, 2.05) | - |
| Other exposed | 111 (29.1) | 7 (19.4) |  | 0.53 (0.18, 1.52) | - |
| Identification Mode |  |  |  |  |  |
| Self-identified | 223 (58.5) | 33 (91.7) | <0.001 | 1.00 | 1.00 |
| Surveillance-identified | 158 (41.5) | 3 (8.3) |  | 0.13 (0.04, 0.43) | 0.17 (0.04, 0.66) |
| Cluster |  |  |  |  |  |
| Scattered | 155 (40.7) | 15 (41.7) | 1.000 | 1.00 | 1.00 |
| Clustered | 226 (59.3) | 21 (58.3) |  | 0.96 (0.48, 1.92) | 2.27 (0.94, 5.47) |

\*AOR, adjusted OR; the model is constructed using the stepwise logistic regression (entry probability 0.05; removal probability 0.10) with variables including age, fever, cough with phlegm, sore throat, headache, breath problem, diarrhea, days from illness onset to the first medical visit, days from the first medical visit to admission, identification mode, and cluster.

## Discussion

In this study, we reported the epidemiological and clinical characteristics of confirmed COVID-19 cases in Shenzhen, a large migrant city with the highest GDP per capita in mainland China. Our results can be used to improve the prediction of transmission risk, design and implementation of intervention measures and strategies, and assessments of intervention effectiveness in similar settings.

The epidemic curve of COVID-19 in Shenzhen had practical implications. Overall, the rapid increase of cases in Shenzhen was interrupted by the massive control measures implemented since Jan 23 and followed by a sustained downward trend. The majority of the cases were imported and the local transmission was limited. Furthermore, most cases were self-identified at the beginning, while cases were more likely to be identified by surveillance afterwards. These changes may be associated with a comprehensive set of interventions including the lockdown of Wuhan and neighboring municipalities since Jan 23, aggressive isolation, screening of high-risk populations, extreme social distancing, and other stringent containment efforts. Our estimated initial reproductive number of 2.5 in early epidemic with rapid growth of cases was similar to estimates of 2.2 to 2.68 elsewhere (*3, 13*). It was speculated that effective reproduction number might have decreased below one recently, given there were no more local cases after Feb 14. Of note, the number of new confirmed COVID-19 cases in Shenzhen continued to drop in recent weeks although businesses had begun to resume since Feb 10. In similar overseas settings where there is high probability of importing COVID-19 cases, the containment success achieved in Shenzhen may be replicated by implementing active mass interventions as early as possible.

Fever and cough were the most common symptoms of COVID-19 on hospital admission in Shenzhen, which was in accordance with findings from a study that included 1099 patients from 30 provinces of China (*14*). This county-wide study also reported that the proportion of patients with fever increased (88.7%) after hospital admission, although the proportion (43.8%) on admission was lower than that in our study (67.4%) (*14*). The overall proportion of patients with cough was lower than that of other study (59.4%) (*5*); meanwhile, more patients had dry cough (34.3%) than cough with phlegm (15.1%) in our study. The main symptoms were significantly different regarding different exposure sources; patients with Shenzhen exposure were less likely to have fever and cough compared to those with Wuhan exposure. In addition, the patients in our study were less likely to have symptoms compared to those patients reported in Wuhan (*15*). Interestingly, surveillance-identified patients showed fewer symptoms at the time of identification than those self-identified patients in our study. This might be because active surveillance identified cases at an early stage of COVID-19.

Our study reported preliminary findings on the clinical severity of COVID-19 in Shenzhen. The proportion of severe or critical cases in Shenzhen (8.6%) was significantly lower than that reported by Guan et al. in patients across China (15.7%) (*15*). In our study, older age, breathing problems, and diarrhea were correlated to the clinical severity on admission. Other studies also suggested that these factors were associated with poorer outcomes in patients in Wuhan and elsewhere (*5, 15*). Furthermore, the longer time from the first medical visit to hospital admission was associated with higher clinical severity on admission, while surveillance identification was associated with lower clinical severity. Thus the low proportion of severe or critical cases in our study might be explained by the large number of cases identified by surveillance and the short time interval from the first medical visit to hospital admission. Of note, 33 (83.3%) of the severe or critical cases were self-identified, which might be due to delayed hospital admission. Through active surveillance efforts such as screening of high-risk populations, a large proportion of COVID-19 patients were identified at the early stage of their illness, thus decreasing the possibility of progression to a severe illness. The hospital fatality rate of COVID-19 in Shenzhen (0.7%) was much lower than 14% reported from Wuhan (*16*), and also lower than 1.4% reported in a county-wide study (*15*). Since data on clinical outcomes were censored in our study, the hospital fatality rate may be underestimated.

Although our study had major strengths such as population-wide case identification in a major migrant city, a large sample size, and complete profiling of epidemics along the timeline of population interventions, there were several limitations that should be acknowledged. First, since some patients were still under treatment so far, we could not yet report the complete data of the disease progression on severity or assess predictors of clinical outcomes. Second, important information, such as timelines of possible exposure, illness onset, and medical visits, was self-reported in the epidemiological investigations, which might be subject to recall bias. Third, we did not have enough information on comorbid conditions,laboratory testing, and radiological examination, which restricted the scope of our analyses. Thus, more detailed clinical characteristics related to COVID-19 could facilitate further analyses in future studies.

In conclusion, our study indicated there was limited local transmission of SARS-CoV-2 in the presence of intensive interventions in Shenzhen, where imported cases accounted for the majority of the confirmed cases. A substantial proportion of the cases were surveillance-identified with less severe illness. Although the intensive and comprehensive measures and interventions have effectively contained the epidemic in Shenzhen, there is still high risk of rebound of COVID-19 cases due to the return of migrants for work, reopening of schools, removal of restrictions of movements and gatherings, and potential importation of cases from other countries. Close monitoring and risk assessments are still imperative for the prevention and control of COVID-19 in the future.

## Data Availability

All data in this manuscript are not available now.

## Acknowledgments

We acknowledge all the staff from the Department of Communicable Diseases Control and Prevention in Shenzhen Center for Disease Control and Prevention. We also acknowledge Joseph Wu, Peng Wu, and Jin Zhao for editing and proofreading of this manuscript.

